# Oropouche virus importation in Southern Brazil and emerging concern calling for enhanced public health surveillance

**DOI:** 10.1101/2025.02.19.25322570

**Authors:** Franciellen Machado dos Santos, Elverson Soares de Melo, Gustavo Barbosa de Lima, Alexandre Freitas da Silva, Marcelo Henrique Santos Paiva, Bartolomeu Acioli-Santos, Clarice Neuenschwander Lins de Moraes, Amanda Pellenz Ruivo, Tatiana Schäffer Gregianini, Milena Bauermann, Thales Bermann, Valeska Lizzi Lagranha, Ludmila Fiorenzano Baethgen, Fernanda Godinho, Gabriel da Luz Wallau, Ana Beatriz Gorini da Veiga, Richard Steiner Salvato

## Abstract

Oropouche virus, an arthropod-borne virus transmitted by *Culicoides paraensis*, is an endemic arbovirus that mostly circulates in the Amazon basin. However, between 2022 and 2024, it reemerged as a more widespread public health concern in South America. We conducted a pooled-sample molecular surveillance to understand the true prevalence of Oropouche fever in Brazil’s southernmost state. Over 18 months, we analyzed 4,060 samples to monitor the virus emergence in the Rio Grande do Sul state. We detected the first human case of Oropouche virus in the state, and our phylogenetic reconstruction indicated a travel-related introduction from Amazon region into Rio Grande do Sul. Despite the absence of local transmission, the invasion of *Culicoides paraensis* and enzootic circulation of the Oropouche virus in Rio Grande do Sul highlight the risk of Oropouche fever outbreaks in the region. We demonstrated that pooled-sample surveillance effectively monitors virus introduction during periods of low endemic circulation, serving as an essential active surveillance tool for the timely detection of virus emergence and enhancing public health preparedness. The multiple introductions of distinct Oropouche virus lineages into southern Brazil underscore the importance of genomic surveillance and public health strategies to monitor and mitigate arbovirus spread in the region.

## 1 Introduction

Oropouche virus (OROV) is a reemerging arthropod-borne virus (arbovirus) primarily transmitted by the bites of *Culicoides paraensis* [1]. The virus is maintained in a zoonotic transmission cycle, including vertebrate hosts such as pale-throated sloths, rodents, non-human primates, and other wild mammals. Oropouche fever is usually characterized by mild symptoms, beginning with the abrupt onset of fever and accompanied by severe headache, chills, myalgia, arthralgia, photophobia, dizziness, retro-orbital pain, nausea, and vomiting. However, in some cases, the disease can progress to more severe manifestations, including hemorrhagic symptoms, meningitis, and meningoencephalitis [1,2]. Recently, complications during pregnancy have been linked to OROV infection, with cases of vertical transmission reported in Brazil [1,3]. These have been associated with adverse pregnancy outcomes, including fetal death and congenital abnormalities [4]. Neither vaccines nor antiviral drugs are available to prevent OROV infection or treat individuals affected by Oropouche fever [1].

The first OROV infections have been documented in the Amazon basin since the 1950s [1]. However, as a typical neglected tropical disease, the true burden of Oropouche fever has remained largely unknown until recently. Current estimates indicate that more than half a million human cases may have occurred since the virus was first identified [1]. In recent years (2022-2024), a significant increase in Oropouche fever cases has been observed in Brazil and other countries such as Bolivia, Colombia, Peru, and Cuba [5]. In Brazil, outbreaks have primarily been reported in the northern states, though different regions outside the endemic areas have also been affected. By February 2025, over 19,000 laboratory-confirmed cases had been reported in Brazil, with almost the totality of the cases reported in the current year occurring in regions considered non-endemic for the virus circulation [6]. A genomic study identified that these outbreaks were caused by a new reassortant virus, which contains genomic segments from different viruses classified within the species *Orthobunyavirus oropoucheense* [7]. As of February 2025, OROV transmission has been identified in 23 of the 27 Brazilian states, except for Rio Grande do Norte, Goiás, Paraná, and Rio Grande do Sul, which have only reported imported cases. To date, four deaths associated with the virus have been confirmed, along with five cases of vertical transmission, including four fetal deaths and one congenital anomaly [8].

Despite the importance of OROV emergence and its rapid expansion across Brazil, routine diagnostic testing for the virus is not widely implemented in arbovirus surveillance conducted by public health laboratories, including Rio Grande do Sul. In light of the emergence of OROV cases in the country, we conducted an active surveillance study to detect the virus in samples from individuals who tested negative for other arboviruses (dengue, Zika, and chikungunya). We aimed to understand better the prevalence of Oropouche fever in the southernmost state of Brazil to guide timely public health interventions.

## 2 Methods

### 2.1 Population

From January 2023 to June 2024, Rio Grande do Sul accounted for 245,000 cases of arboviruses infections, almost the totality of these cases due to dengue virus infection. In the study period, the Rio Grande do Sul Public Health Laboratory (LACEN-RS) received 62,700 samples from cases suspected of infection by arboviruses. These samples were analyzed using antigen tests to detect the dengue virus’s non-structural protein 1 (NS1), immunoglobulin (Ig)M antibodies and RT-PCR for dengue, zika, and chikungunya detection. Of the samples, 37,700 were collected within the first five days of symptom onset. We then performed a probabilistic random selection, considering the collection date and city, to choose 4,060 samples collected within five days of symptom onset, with negative results for the tested viruses (dengue, zika, and chikungunya) in routine surveillance.

### 2.2 Pooled-sample Oropouche virus detection

The 4,060 samples were pooled into 406 pools, with 10 samples per pool. These pools were formed by grouping samples from the same geographic region (e.g., city) and with similar collection times to achieve geographic and temporal closeness. For pooling the samples, 50 µL of serum from each of the 10 samples in the same pool were combined in a microtube, resulting in a total volume of 500 µL. Subsequently, 200 µL of this volume was used for nucleic acid extraction. The extraction was performed on Extracta-96 equipment (Loccus, São Paulo, Brazil) using the Fast Kit–Viral DNA and RNA (MVXA-P096 FAST), according to the manufacturer’s instructions. The OROV detection was performed through reverse transcription-quantitative polymerase chain reaction (RT-qPCR) using a previously published protocol [9] and performed in a CFX Opus Real-Time PCR System (Bio-Rad, Hercules, CA, USA).

### 2.3 Viral sequencing and genome assembly

OROV genome amplification and sequencing library was prepared using the Illumina COVIDSeq Kit (Illumina, San Diego, USA) replacing the SARS-CoV-2 primers by OROV previously designed [7]. Genome sequencing was conducted on the MiSeq platform with the V2 reagent kit, employing a paired-end approach with 2× 150 cycles. The resulting sequencing reads were processed using the ViralFlow v.1.2.0 pipeline [10], applying a reference-guided genome assembly strategy. Each of the three genomic segments of OROV—L (OL689334.1), M (OL689333.1), and S (OL689332.1)—was assembled independently using the corresponding reference sequences.

### 2.4 Phylogenetic analysis

To determine the context of the OROV sample collected in the Rio Grande do Sul in the virus’s spread across Brazil, we aligned the genome sequenced in this study along representative OROV genomes to contextualize the phylogenetic relationships of the strain, its transmission dynamics and potential epidemiological implications. We included OROV genomes from Gräf et al. (2024) [11] and Naveca et al. (2024) [7], as well as additional genomic sequences from samples available in the GISAID database (Global Data Science Initiative—an essential platform for sharing viral genomic data (WALLAU et al., 2023) [12] through the EpiArbo database, retrieved up to January 10, 2025. Concatenated OROV segments were aligned using MAFFT (version 7.490) [13], followed by manual curation to remove unaligned regions and non-coding sequences.

The alignment of concatenated segments was analyzed using ModelFinder [13,14] to decide the most appropriate nucleotide substitution model. The sequences were then processed in BEAST (version 1.10.4) [15] to reconstruct a Bayesian time-scaled phylogenetic tree. Sample collection dates were incorporated as Tip Dates. The analysis was conducted under the GTR+G+I substitution model, with a partitioning scheme that treated each codon position separately. An uncorrelated lognormal relaxed clock model was also applied. As a tree prior, the coalescent Bayesian Skyline model was used, enabling the inference of changes in adequate population size over time without imposing strict parametric assumptions on viral demography. Ten independent runs were performed using the Markov Chain Monte Carlo (MCMC) for the phylogenetic inference method, each consisting of 100 million generations. The chains from all runs were combined using LogCombiner (part of the BEAST package), discarding the first 10% of states as burn-in. Chain convergence was assessed by calculating the Effective Sample Size (ESS) for all parameters using Tracer (version 1.7.1) [16], ensuring values above 200 for a robust estimation of posterior distributions. The consensus phylogenetic tree was obtained using the maximum clade credibility criterion with TreeAnnotator (part of the BEAST package) and visualized and annotated in FigTree.

## 3 Results

In 2023, Brazil reported a total of 832 cases of Oropouche fever, with the majority of these cases concentrated in the endemic Amazon Basin region. In 2024, the number of Oropouche fever cases surged to 13,783, marking a clear and concerning spread of OROV across all Brazilian regions. By 2025, the pattern persisted, with 4,857 cases reported so far, the majority of which occurred outside the historically endemic Amazon Basin region, highlighting a significant shift in the epidemiology of the disease (Figure 1A).

**Figure 1.**
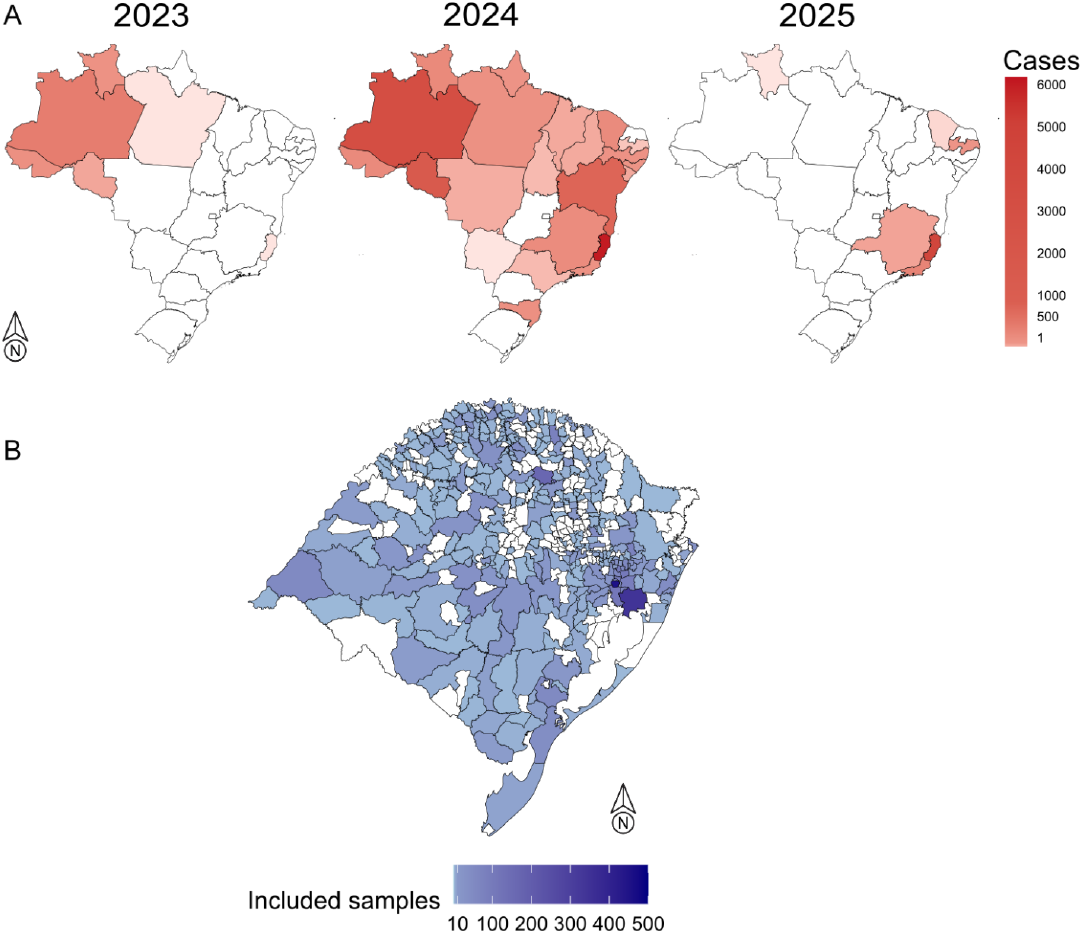
Distributions of confirmed Oropouche fever cases reported in Brazil from 2023 to 2025 and geographic distribution of the samples included in this study. (A) Number of confirmed Oropouche fever cases reported in Brazilian states per year, including data up to February 18th, 2025, as reported by the Brazilian Ministry of Health. (B) The map of Rio Grande do Sul state, showing the geographic distribution of clinical samples from individuals presenting arboviruses infection symptoms, tested for OROV infection in this study.

Among the 4,060 samples tested (Figure 1B), a single positive case for OROV was identified. Initially, one of the tested pools was positive for OROV, with amplification observed at a cycle threshold (Ct) 17. Subsequently, RNA was extracted from the ten individual samples within this positive pool and subjected to RT-PCR. Amplification was detected in one of these samples, which yielded a Ct value of 16. The sample was obtained from an individual diagnosed in Erechim, located in the northern region of Rio Grande do Sul state. The patient, a resident of the bordering municipality of Aratiba, presented with symptoms including fever, myalgia, and a rash on January 8th, 2024. The NS1 test to detect the non-structural protein 1 of the dengue virus was negative, while RT-PCR confirmed the presence of OROV. However, the case was considered likely imported, as the individual had traveled to the northern region of Brazil one week before the onset of symptoms. To investigate the hypothesis of travel-related importation, we sequenced the OROV genome using an amplicon-based sequencing protocol, and performed a phylogenomic analysis along with OROV genome sequences available in the GISAID database.

The reconstructed time-scaled phylogeny of the virus’s evolution in Brazil (Figure 2A) indicates that the OROV sample isolated in Erechim clusters with sequences from the state of Amazonas, specifically within the AM-I clade. This clade comprises isolates from the central region of Amazonas, where the virus circulated actively between November 2023 and February 2024, coinciding with the reported travel period of the patient to the region. In addition to this travel-related viral introduction into Rio Grande do Sul, we identified that available genomes from Paraná and Santa Catarina, two other southern Brazilian states, did not cluster with the genome from patient diagnosed in Rio Grande do Sul, further supporting the travel-related infection. This data reinforces that the case analyzed in this study did not result from a viral importation from the neighboring states.

**Figure 2.**
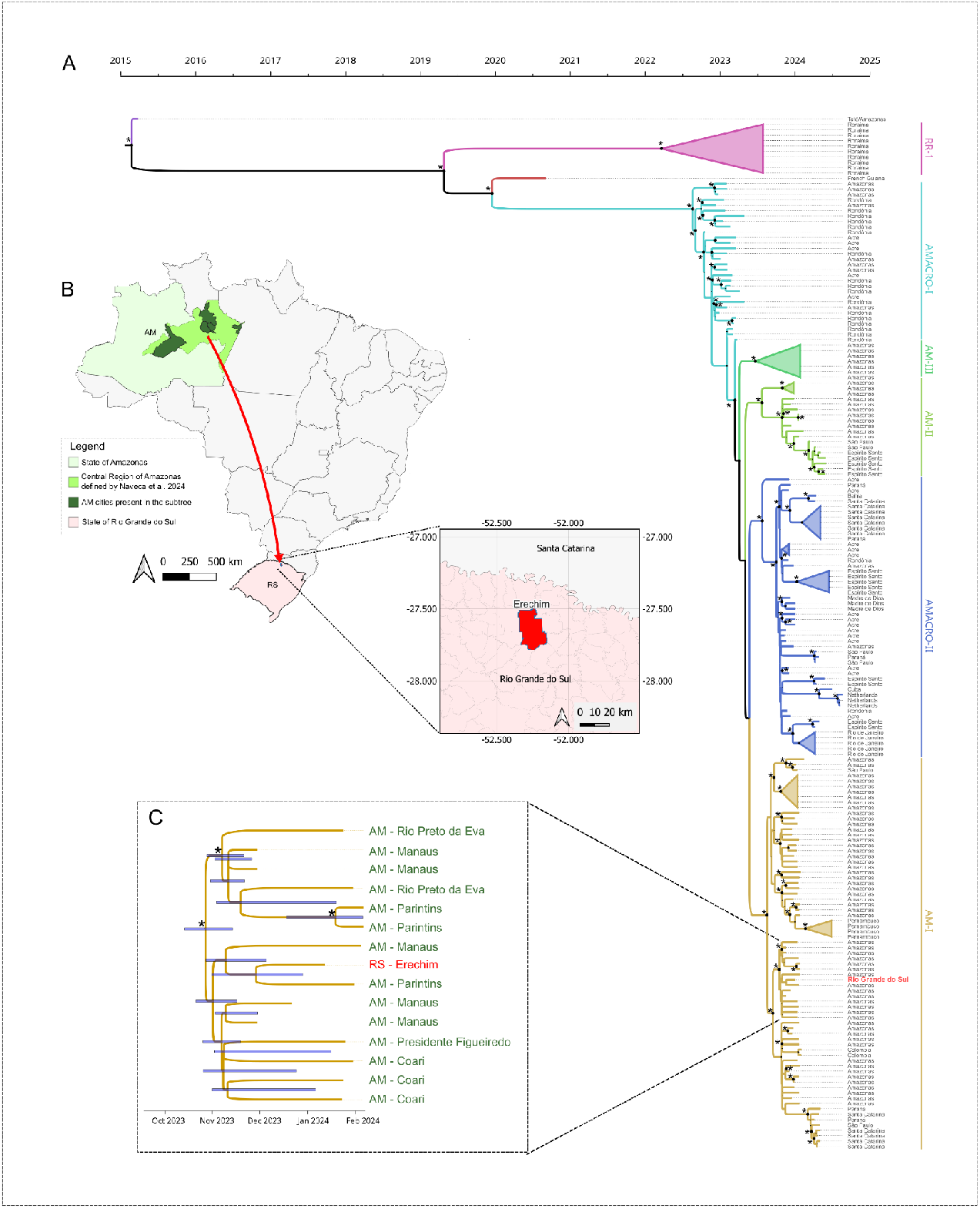
Phylogenetic analysis of Oropouche virus (OROV) in Brazil, highlighting a sample from Rio Grande do Sul. (A) Maximum Clade Credibility (MCC) time-scaled phylogenetic tree showing six major OROV clades defined by Naveca et al. (2024). The Rio Grande do Sul sample (red) clusters within the AM-I lineage from Amazonas. A subtree within this well-supported clade (posterior probability = 1) includes sequences from Erechim (Rio Grande do Sul) alongside five Amazonas cities. The Most Recent Common Ancestor (MRCA) is estimated to have emerged between mid-October and mid-November 2023. The map in (B) shows the transmission route from the central region of Amazonas to Erechim). The asterisks in the left side of nodes in (A) and C represent posterior probabilities higher than 0.9. In A, these probabilities are visually represented by gradually sized points at the tree’s nodes.

A more detailed analysis of the highly supported subclade where the Erechim sample is positioned (Figure 2C), indicates that this virus shares a recent common ancestor with samples from central Amazonas (highlighted in light green in Figure 2B), including ones from Rio Preto da Eva, Parintins, Presidente Figueiredo, Coari, and Manaus cities. The estimated time to the most recent common ancestor (TMRCA) suggests that this subclade emerged between mid-October and mid-November 2023, coinciding with the peak circulation of the AM-I clade in the central region of Amazonas, occurring just one month prior to the travel of the individual from the positive case in Erechim to Manaus, Amazonas. These findings, in addition to the travel history of this individual diagnosed with Oropouche fever, support the hypothesis that the infection was acquired in the Central Region of Amazonas and subsequently introduced directly into Erechim, as illustrated in Figure 2B.

## 4 Discussion

During the study period, Brazil faced a significant OROV outbreak, marked by the virus’s spread from the Amazon Basin region to other parts of the country. This was accompanied by a notable shift in the epidemiology of Oropouche fever, with increasing cases now emerging in the Southeast of Brazil. As of February 2025, 23 out of Brazil’s 27 states had reported cases of Oropouche fever, highlighting the expanding spread of the virus across the country. Notably, Rio Grande do Sul, the southernmost Brazilian state, did not register any cases during this period. We employed pooled-sample testing to enhance surveillance and investigate the potential introduction of the OROV in the region. Out of the 4,060 samples tested, we identified only one positive sample for OROV, demonstrating the ability of pooled surveillance testing to detect infections even in scenarios of low viral circulation.

Pooled-sample surveillance has emerged as a highly effective strategy for enhancing the effiiciency of clinical diagnostics [17]. By significantly reducing the time, labor, and reagents required for large-scale testing, this method enables effiicient screening for pathogens of large amounts of asymptomatic populations. Its simplicity, alignment with existing approved procedures, and lack of need for specialized sample handling or additional information make it an easily scalable solution. These advantages establish pooled-sample surveillance as a practical and adaptable tool for public health screening, particularly in scenarios demanding rapid and widespread testing, such as epidemics or outbreaks [18–20]. However, most reported implementations of this approach focus on SARS-CoV-2, and there is limited evidence demonstrating the use of pooled surveillance testing for arbovirus detection.

The time-scaled phylogeny of the detected OROV revealed critical patterns of viral spread, particularly between northern and southern regions. The sample detected in Erechim, Rio Grande do Sul, clustered within the AM-I clade, linking it to active viral circulation in central Amazonas between late 2023 and early 2024. The estimated emergence of this subclade in mid-October to mid-November 2023, coupled with the patient’s travel history to Manaus, supports the hypothesis of direct viral introduction from Amazonas to Rio Grande do Sul. Furthermore, distinct introductions into neighboring Santa Catarina and Paraná states, represented in separate clades, further support the hypothesis of viral importation from the Amazon region rather than local transmission across bordering states. These findings emphasize the crucial role of genomic surveillance in monitoring arbovirus spread, especially in areas with high human mobility and ecological diversity. Identifying multiple independent introductions into southern Brazil stresses the need for enhanced public health strategies to monitor and mitigate the spread of emerging arboviruses [12].

In the first months of 2024 was detected the infestation by *Culicoides paraensis*, the main OROV vector, in seven municipalities from Rio Grande do Sul state (Mampituba, Três Forquilhas, Dom Pedro de Alcântara, Itati, Maquiné, Mampituba, and Terra de Areia). This detection was due to the entomological surveillance activities conducted by the State Center for Health Surveillance and the Department of Agriculture, Livestock, Sustainable Production, and Irrigation of Rio Grande do Sul [21]. Such findings reveal a concerning scenario for the potential transmission of the virus, as the presence of *Culicoides paraensis* in the region creates an environment conducive to the spread of the virus. Aside from the invasion of the biting midge, previous studies conducted in the area have also reported the enzootic circulation of OROV, evidenced by antibodies in non-human primates in 2004, 2012, and 2014 [22]. This combination of vector presence and historical evidence of viral circulation increases the risk of Oropouche fever outbreaks in the region, given the potential for both vector expansion and ongoing enzootic circulation.

In recent years, Rio Grande do Sul has experienced a change in dengue occurrence patterns, from self-limited transmission due to virus importation events to annual large dengue virus outbreaks by lineages that are now endemic in the region. This change in the dengue transmission pattern led to significant morbidity and mortality increased burden, as the Rio Grande do Sul had a naive population for dengue infection. This epidemiological change is mainly associated with *Aedes aegypti* (the primary dengue mosquito vector), which spread to southern areas due to the erosion of the climatic barrier related to climate change, leading to new outbreaks in an immunologically naive human population [23].

A similar scenario is being remounted in Rio Grande do Sul, where an entirely naive population for OROV is present. As evidenced by entomological surveillance data, the region has witnessed an invasion of *Culicoides paraensis*, and considering the current climate change scenarios, the dispersion of this vector is expected to expand. Additionally, the increased viral fitness of the OROV reassortant may drive its spread and facilitate its expansion into previously non-endemic areas, such as Southern Brazil [24]. This complex scenario of rapid viral expansion, the spread of *Culicoides paraensis*, and the current lack of Oropouche surveillance in south Brazil presents a significant public health threat, leaving the region highly vulnerable to potential outbreaks.

The occurrence of a potential OROV outbreak in Southern Brazil, along with the increasing annual outbreaks of dengue virus occurring in recent years, could dangerously increase the disease burden in the region, leading to overcrowding in healthcare services, increased demand for resources such as medical supplies and healthcare professionals, resulting in significant social and economic impact. Active surveillance allows the early detection of viral introduction and spread, enabling the timely implementation of disease control measures such as vector mosquito control, health education campaigns, and healthcare system preparedness. This proactive approach is crucial in preventing a health crisis and averting the collapse of medical services.

Our study demonstrated that during periods of low endemic circulation, sentinel laboratory surveillance using a pooled sampling approach can serve as an effective alternative for monitoring the potential introduction of new pathogens. This method proves valuable in settings already burdened with other diseases, such as dengue, by providing effiicient surveillance without overwhelming existing routines.

## Data Availability

All data produced in the present study are available upon reasonable request to the authors.

## Ethics Approval

This project was approved by the Research Ethics Committee (CEP) at Escola de Saúde Pública (SES-RS). Process number: CAAE: 67181123.1.0000.5312

## Acknowledgments

We gratefully acknowledge all data contributors, i.e., the Authors and their originating laboratories responsible for obtaining the specimens and their submitting laboratories for generating the genetic sequence and metadata and sharing via the GISAID Initiative, on which this research is based.

